# Identification of the immune-associated enhancer RNA SATB1-AS1 as a novel biomarker for thymic cancer prognosis

**DOI:** 10.1101/2025.02.13.25322204

**Authors:** Hongpeng Wang, Yichu Huang, Guang-tao Min, Lei Jiang

**Affiliations:** Department of Colorectal Surgery, First Clinical Hospital of Lanzhou University, Lanzhou, Gansu, P.R. China; The First Clinical Medical College of Lanzhou University, Lanzhou, Gansu, P.R. China

**Author notes:** **corresponding author Name:** Lei Jiang, **Department:** Department of Colorectal Surgery, First Clinical Hospital of Lanzhou University, Lanzhou, Gansu, P.R. China, **Email address:**, **Name:** Guang-tao Min, **Department:** Department of Colorectal Surgery, First Clinical Hospital of Lanzhou University, Lanzhou, Gansu, P.R. China, **Email address:**. **Name:** Hongpeng Wang, **Department:** Department of Colorectal Surgery, First Clinical Hospital of Lanzhou University, Lanzhou, Gansu, P.R. China, **Email address:**. **Name:** Yichu Huang, **Department:** The First Clinical Medical College of Lanzhou University, Lanzhou, Gansu, P.R. China. **Name:** Guang-tao Min, **Department:** Department of Colorectal Surgery, First Clinical Hospital of Lanzhou University, Lanzhou, Gansu, P.R. China, **Email address:**.

## Abstract

**Objective:** To screen key enhancer RNAs (eRNAs) in thymoma (THYM) through The Cancer Genome Atlas (TCGA) database and explore their potential as prognostic molecules and therapeutic targets in THYM.

**Methods:** Gene expression RNA-seq profiles of 33 cancer types were retrieved and downloaded from the TCGA database, and Kaplan–Meier survival analysis and Spearman correlation analysis were applied to screen eRNAs and target genes associated with survival in THYM patients. The correlation of target eRNAs with clinical features was assessed. Gene set enrichment analysis was performed to investigate the potential biological functions of target eRNAs. Validation of the prognostic potential of target eRNAs across cancers in other cancer types.

**Results:** In THYM, SATB1-AS1 and the target gene SATB1 were associated with poor patient prognosis, and SATB1-AS1 expression was correlated with patient stage (P<0.05). Gene and pathway enrichment analyses revealed that SATB1-AS1 was associated with leukocyte transendothelial migration, natural killer cell-mediated cytotoxicity, neutrophil extracellular trap formation, PD-L1 expression and the PD-1 checkpoint pathway in cancer.

**Conclusion:** SATB1-AS1 may be a key eRNA in THYM and has the potential to be a marker and therapeutic target for the early diagnosis and prognosis of THYM.

## 1 Introduction

Thymoma (THYM) is the most common tumor type in the anterior mediastinum, accounting for approximately 95% of all thymic tumors, with an incidence rate of 4.09 per million in China, which is much higher than that in other countries in the world, including Europe[1]. Milan Radovich et al. systematically classified thymomas and reported that histologic subtypes A, AB, B1, B2, B3, and thymic carcinoma (TC) were significantly associated with multiple gene mutations[2]. Notably, among many advanced tumors, the 5-year median survival rate for thymomas is 69%, whereas the 5-year median survival rate for TC is only 36%[3]. Surgery is the treatment of choice for tumor management. Surgery, as the treatment of choice for tumor therapy, is the only choice for patients with early-stage thymoma, whereas palliative treatments such as radiotherapy have become the only option for patients with advanced unresectable tumors[4]. In recent years, the emergence of immunotherapy and targeted therapy has provided many new insights into the treatment of patients with thymomas and has also shown some therapeutic promise[5]; however, further development of more individualized treatment regimens to increase disease responsiveness is still needed.

Enhancers are DNA elements that bind to cofactors and transcription factors (TFs), are located in noncoding regions of the genome, have a characteristic chromatin structure, and are involved in driving the transcription of target genes in a variety of ways, such as three-dimensional chromatin remodeling and enhancer–promoter loops[6]. It is also involved in driving the transcription of target genes in various ways, such as three-dimensional chromatin remodeling and enhancer–promoter loops. With the development and advancement of genome sequencing and large-scale genome-wide association studies (GWASs), it has been demonstrated that active enhancers can transcribe noncoding RNAs (ncRNAs) called enhancer RNAs (eRNAs) and that eRNA-mediated enhancer activity plays a definitive role in the progression of many cancers[1]. It has been shown that lncRNAs (NR2F1-AS1, LINC00665, and RP11-285A1.1) and miRNAs (hsa-miRNA-143, hsa-miRNA-141, hsa-miRNA-140, and hsa-miRNA-3199) are significantly correlated with prognosis and overall survival in THYM[7]. Genetic variation in eRNA transcripts was analyzed in more than 30 different types of cancers from TCGA via eRNA quantitative trait loci (eRNAQTLs), confirming that the activation of enhancers and the transcription of eRNAs usually induce mutations in oncogenes or the activation of oncogenic signaling pathways[8]. A recent study also revealed that eRNA transcripts are not associated with oncogenic genes. A recent study revealed that eRNA (CRISPR-Cas9) can control the oncogenic activity of the MYB and DCTD genes and is a key determinant of B-cell precursor acute lymphoblastic leukemia (B-ALL) [9].

However, studies on eRNAs are still limited, and the functions and roles of most eRNAs are still unclear. THYM-associated eRNA regulatory mechanisms are largely worth exploring. Here, we aimed to screen prognostic eRNAs and their target genes in THYM. We found that the eRNA SATB1-AS1 was significantly associated with the survival of SCCHN patients through regulating SATB1 and was correlated with the local immune environment of ESCA, which is expected to be a new biomarker for THYM prognosis and immunotherapy response.

## 2 Materials and methods

### 2.1 Acquisition of the THYM of eRNA data

The Cancer Genome Atlas (TCGA) provides most of the expression data of cancer patients and their clinical data.[10] We first screened 139 THYM-related patients via the TCGA website and downloaded their transcript data. Next, the transcript IDs were converted to gene symbol IDs via the pl language and differentiated between tumor patients and controls. We also used the UCSC Xena online database (https://xena.ucsc.edu/) to obtain clinical data and expression profiling data for 33 cancers. The study was conducted in accordance with the Declaration of Helsinki (revised 2013).

### 2.2 Screening for THYM-related differential eRNAs

The R package “limma” was used to screen for differentially expressed eRNAs[11]. The R package “limma” was used to screen for differentially expressed eRNAs, and patients were categorized into low- and high-expression groups on the basis of the median expression value of each eRNA. The R packages “survival” and “survminer” were used to construct a loop function to perform a log-rank test for each eRNA and plot the Kaplan–Meier survival curve[12]. P<0.05 was considered statistically significant.

### 2.3 Identification of THYM coexpressed genes and gene set enrichment analysis of prognostically relevant eRNAs

Genes with correlation coefficients R>0.7 and P<0.001 were considered coexpressed genes of prognostically associated eRNAs. Functional enrichment analysis of the coexpressed genes revealed their biological functions in THYM. Gene Ontology (GO) and Kyoto Encyclopedia of Genes and Genomes (KEGG) enrichment analyses of coexpressed genes for prognosis-associated eRNAs were performed via the R software package “clusterProfiler” and then visualized via the online analysis website bioinformatics.

### 2.4 Analysis of common mutations of SATB1 in THYM

The TCGA database provides exome sequencing data of nearly 30 tumors, and we classified 119 samples with mutations into two groups according to the expression of SATB1, the target gene of SATB1-AS1, and compared the mutation frequency of related genes between the two groups to calculate the tumor mutation burden (TMB)[13]. The R package “maftools” was used for cluster analysis, and finally, the R package “waterfall” was used to draw waterfall plots to visualize the data.

### 2.5 Correlation analysis of SATB1-AS1 immune checkpoints and immunoregulatory genes

For the correlation analysis of immune-related genes, we extracted the ENSG00000182568 (SATB1) gene and 60 genes of two types of immune checkpoint pathways (inhibitory (24), stimulatory (36) and 150 genes of five types of immune pathways (chemokine (41), receptor (18), MHC (21), immunoinhibitor (24), and immunostimulator (46))[14]. The results were filtered out by screening all normal samples, and a log2 (x+0.001) transformation was applied to each expression value, which ultimately resulted in a sparse correlation between SATB1 and immunomodulatory genes in a variety of cancers.

### 2.6 SATB1-AS1 pancancer survival and correlation analysis

Patients with 32 other cancers were categorized into low and high SATB1-AS1 expression groups according to their median SATB1-AS1 expression values via the R package “limma”. The prognostic value of SATB1-AS1 was investigated via the Kaplan–Meier method, and the correlation between SATB1-AS1 and pancancer was evaluated via the Spearman correlation test.

### 2.7 Statistical analysis

The data were statistically analyzed via R software (v4.4.1). Spearman correlation was applied to estimate the strength of correlation between two samples. Survival analysis was performed via the Kaplan–Meier method, and comparisons between clinical variables in the two groups were performed via the Wilcoxon rank sum test. p<0.05 was considered statistically significant.

## 3 Results

### 3.1 Screening for eRNAs associated with THYM survival

Patient eRNA expression data and clinical survival-related information were combined, and the data were normalized (log2 (x+0.001)), while information on patients with eRNA expression of 0 and normal controls was removed. With eRNA expression divided into high and low groups according to the median expression value, 133 eRNAs significantly associated with OS were identified via the Kaplan–Meier log-rank test (P<0.05) (Supplementary Table S1). After further screening, only 10 genes were significantly correlated with the predicted mRNA levels of the target genes (coefficient of r > 0.7, P<0.001), as shown in Figure 1.

**Fig. 1.**
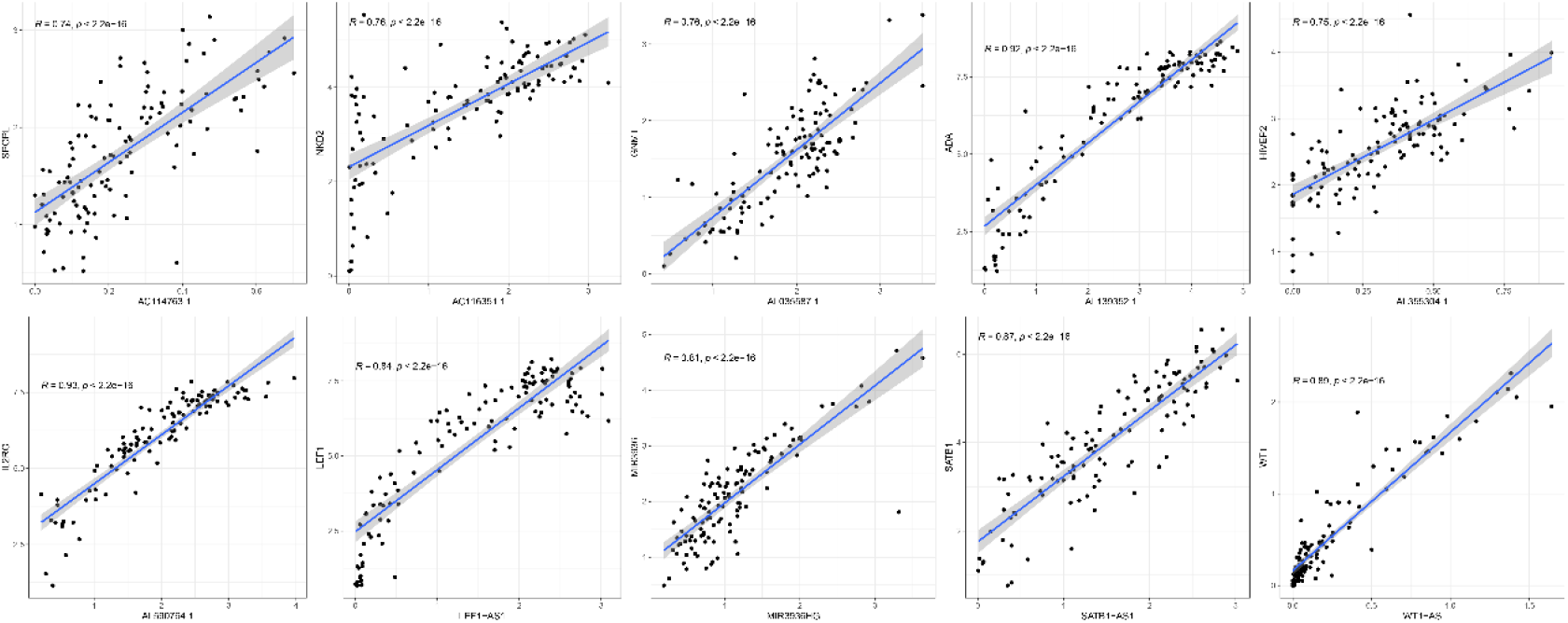
Scatterplot of correlation analysis showing 10 eRNAs strongly associated with THYM survival.

### 3.2 The eRNA SATB1-as1 had the highest positive correlation with its target gene SATB1 in THYM prognosis

Among the 10 prognostically relevant eRNAs, SATB1-as1 had the highest prognostic correlation with THYM (Figure 2A). Therefore, SATB1-AS1 was identified from the 10 eRNAs in this study. We found that high SATB1-AS1 expression was associated with a favorable prognosis in OV patients. In addition, higher SATB1-AS1 expression was more strongly associated with tumor grade and lymphatic infiltration than lower SATB1-AS1 expression was, and lower SATB1-AS1 expression was more strongly associated with tumor grade and lymphatic infiltration. However, we also found that SATB1-AS1 expression levels were independent of age, sex, and tumor grade. These results suggest that SATB1-AS1 may be a favorable independent prognostic biomarker for THYM patients.

**Fig. 2.**
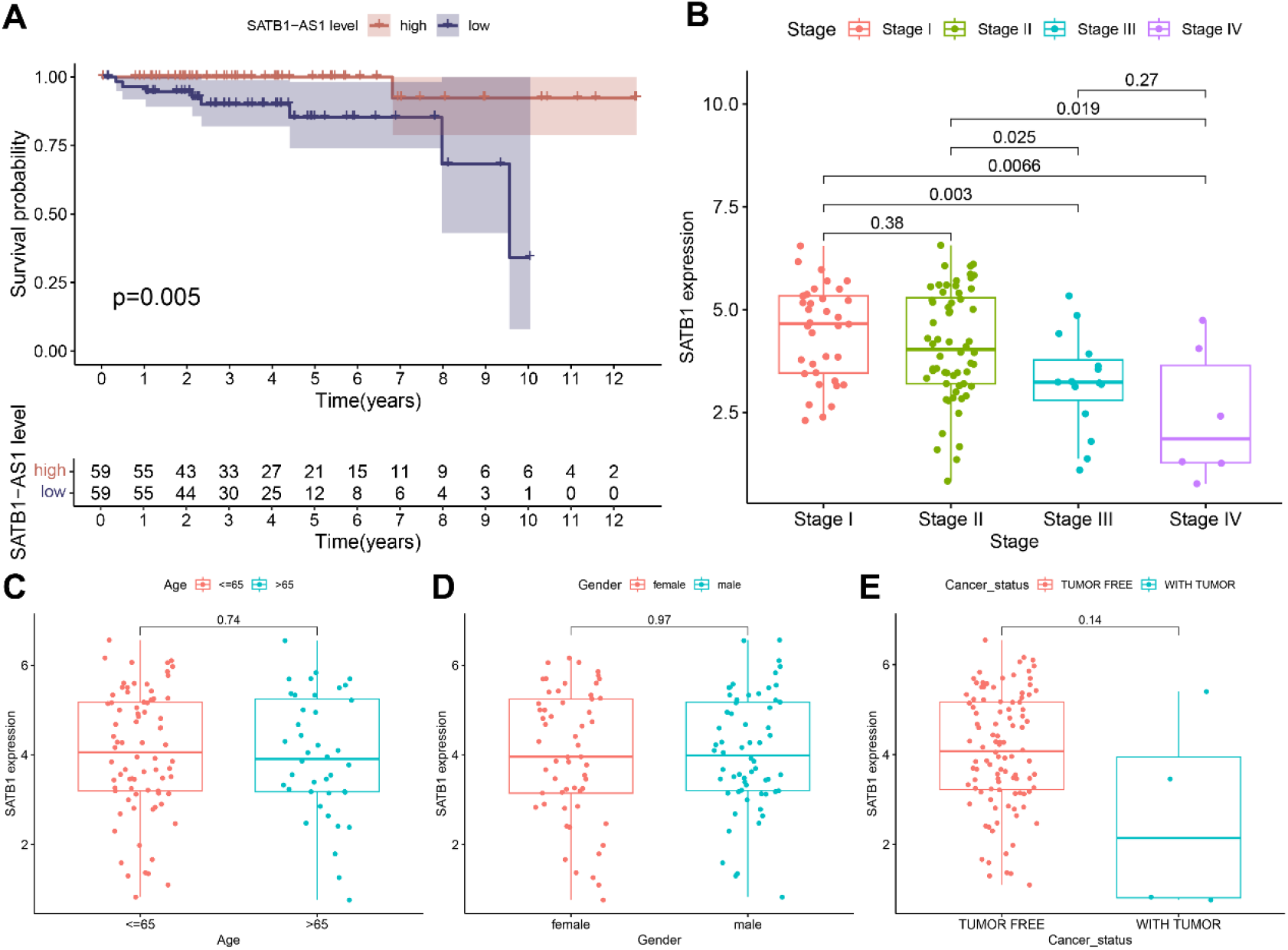
The clinical relevance of eRNA SATB1-AS1 to THYM. **A** Kaplan–Meier OS curve for THYM patients with SATB1-AS1-high and SATB1-AS1-low expression. **B-E** Clinical relevance of eRNA SATB1-AS1 expression in THYM patients.

### 3.3 SATB1-AS 1 is associated with multiple immune-related signaling pathways

GO enrichment revealed that in the BP category, SATB1-AS1 was associated with chromosome segregation, nuclear division and organelle fission (Fig. 3A, B). In CC, SATB1-AS1 is involved mainly in the T-cell receptor complex, plasme membrane signaling receptor complex and chromosomal region.

**Fig. 3.**
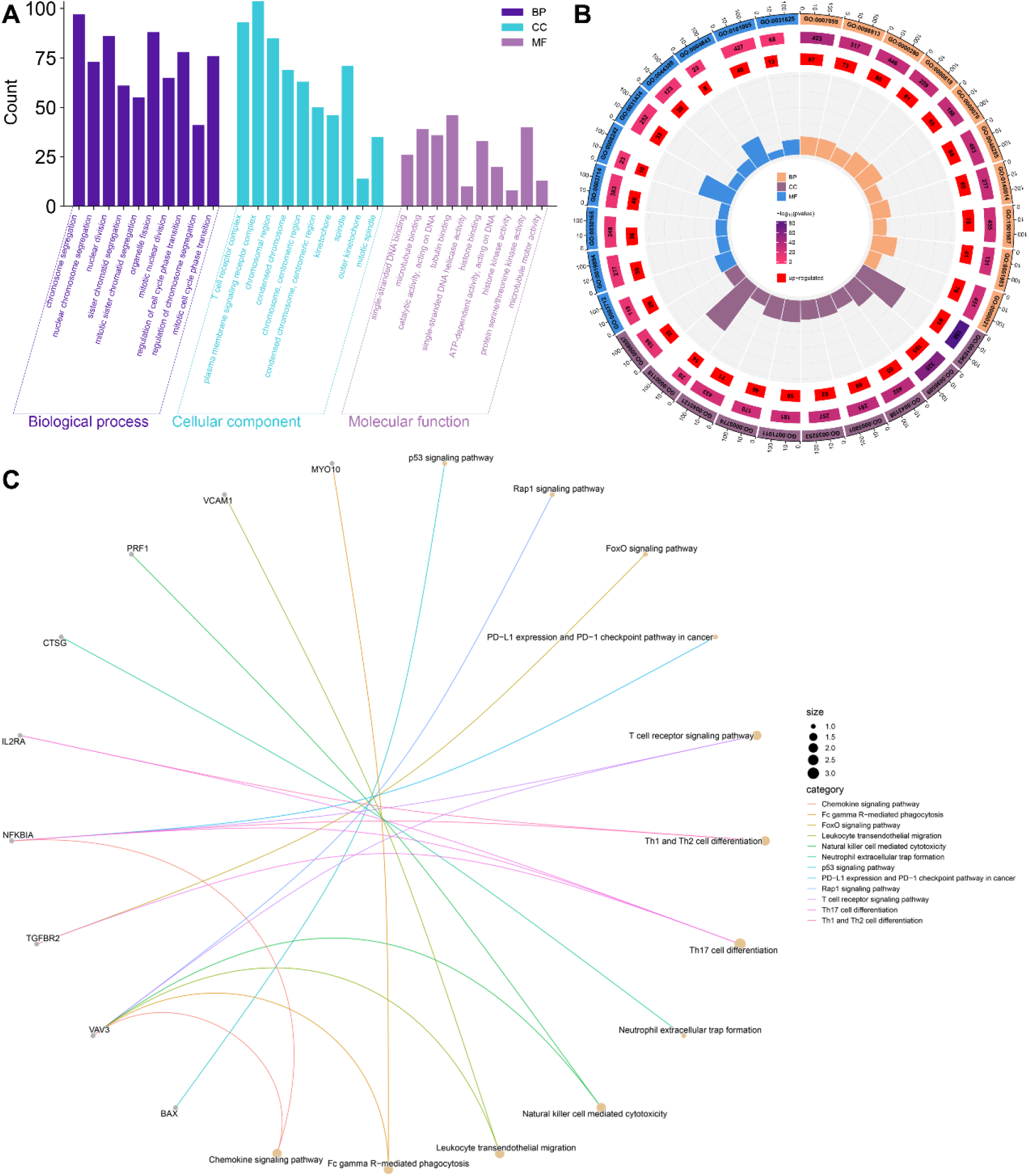
Significant GO and KEGG pathway analysis of SATB1-AS1 co-expressed genes. **A-B**: GO enrichment analyses of the top 10 terms in biological processes, cellular components and molecular functions; **C**: KEGG pathway analyses of the top 12 terms in species enrichment.

In MF, SATB1-AS1 was involved mainly in microtubule binding, protein serine/threonine kinase activity and tubulin binding. The KEGG signaling pathway was also enriched in several signaling pathways related to immune cells, e.g., leukocyte transendothelial migration, natural killer cell-mediated cytotoxicity, neutrophil extracellular trap formation, PD-L1 expression and the PD-1 checkpoint pathway in cancer, the T-cell receptor signaling pathway, Th1 and Th2 cell differentiation and Th17 cell differentiation. Thus, we performed a Spearman correlation analysis between SATB1, the target gene of SATB1-AS1, and immune-related genes and found that the expression of SATB1 was significantly associated with several immune checkpoint marker genes as well as immune immunization pathway-related genes (Figure 4A, B).

**Fig. 4.**
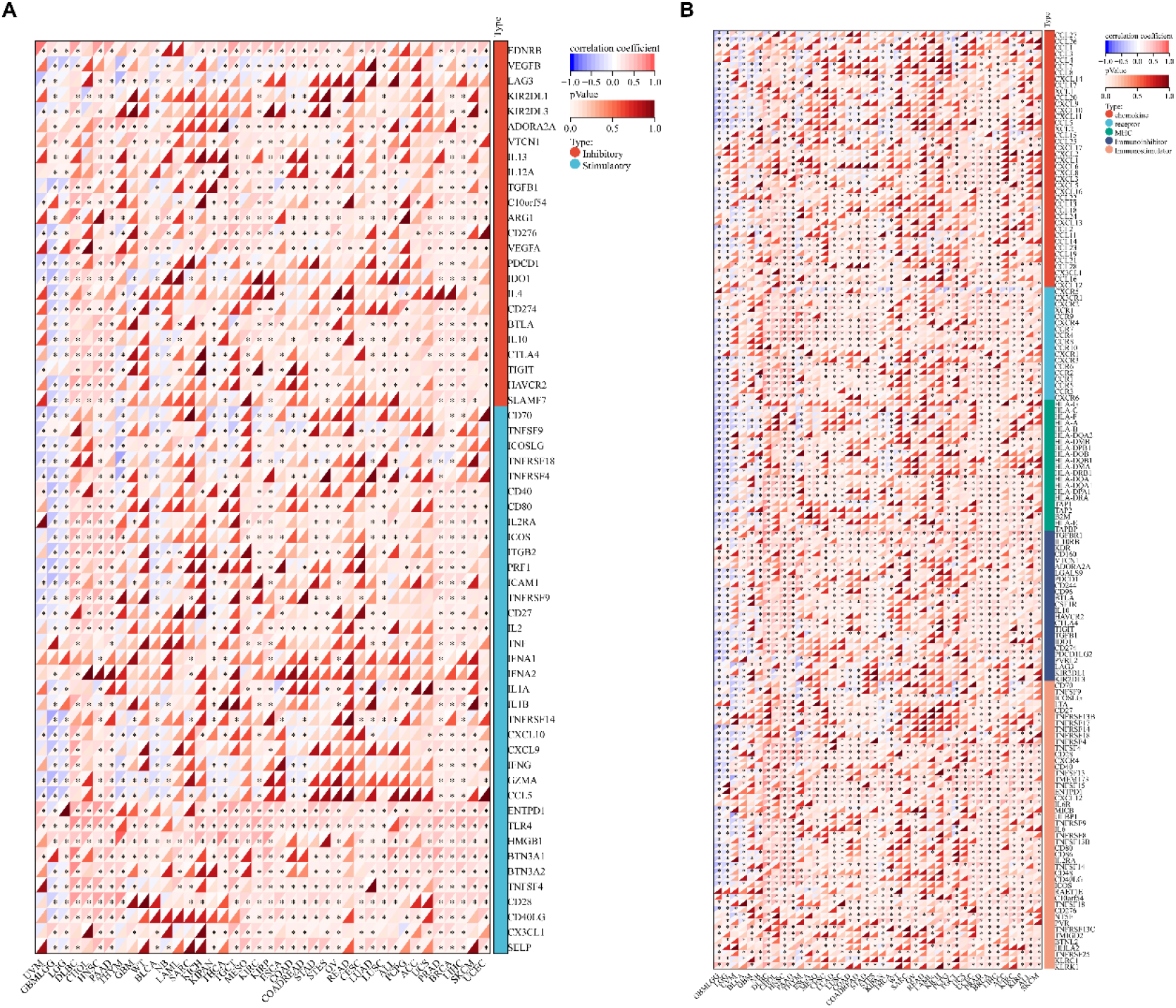
SATB1-AS1 co-expressed genes and immune genes between several cancers in spearman correlation. **A** SATB1 gene was significantly correlated with 60 two-class immune checkpoint pathway genes (Inhibitory(24), Stimulatory(36). **B** SATB1 gene was significantly correlated with 150 five-class immune pathways (chemokine (41), receptor(18), MHC(21), Immunoinhibitor(24) and Immunostimulator(46)) marker genes.

### 3.4 The mutational landscape of SATB1

TMB could serve as an emerging tumor immunotherapy biomarker[15], so we assessed the differences in the frequency of mutations in each set of samples via chi-square tests. In THYM, the most relevant and frequent missense mutations associated with SATB1 were those in GTF2I and HRAS (Figure 5A). Figure 5B shows the relationship between SATB1 mRNA expression and its putative copy number alterations (CNAs).

**Fig. 5.**
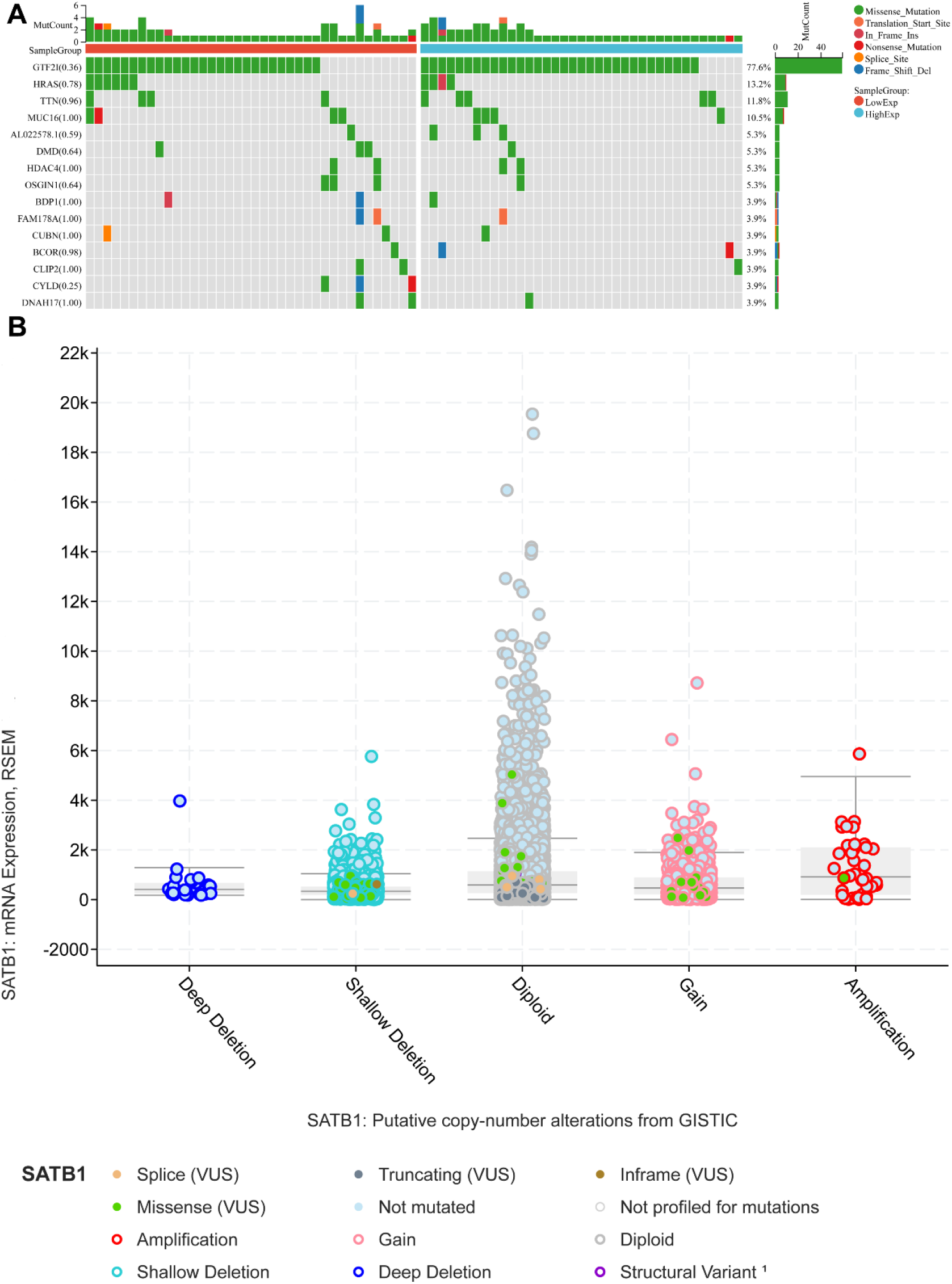
Mutational landscape of SATB1-AS1 in thymic carcinoma. **A** SATB1-AS1 co-expressed genes in THYM Tumor mutation burden (TMB) per sample. **B** The study of the origin of SATB1 in many TCGA cancers by the cBioPortal database

### 3.5 Pan-cancer analysis of SATB-AS1

To assess the tissue specificity of SATB1-AS1, we calculated the difference in expression between normal and tumor samples in each tumor via R software and analyzed the significance of the difference via unpaired Wilcoxon rank sum and signed rank tests (Figure 6A). We observed significant upregulation in seven tumors, such as GBMLGG, LGG, WT, ALL, LAML, PCPG tumors and KICH, and we observed significant downregulation in 25 tumors, such as GBM, KIRC, LUSC, LIHC, SKCM, BLCA, THCA, OV, PAAD, TGCT, UCS, and ACC. Survival analysis of SATB1-AS1 in cancers other than THYM revealed that SATB1-AS1 also had a significant effect on survival in BLCA, KIRC, LAML, LGG, LUAD, SARC, SKCM and UVM (p< 0.05, Figure 6B).

**Fig. 6.**
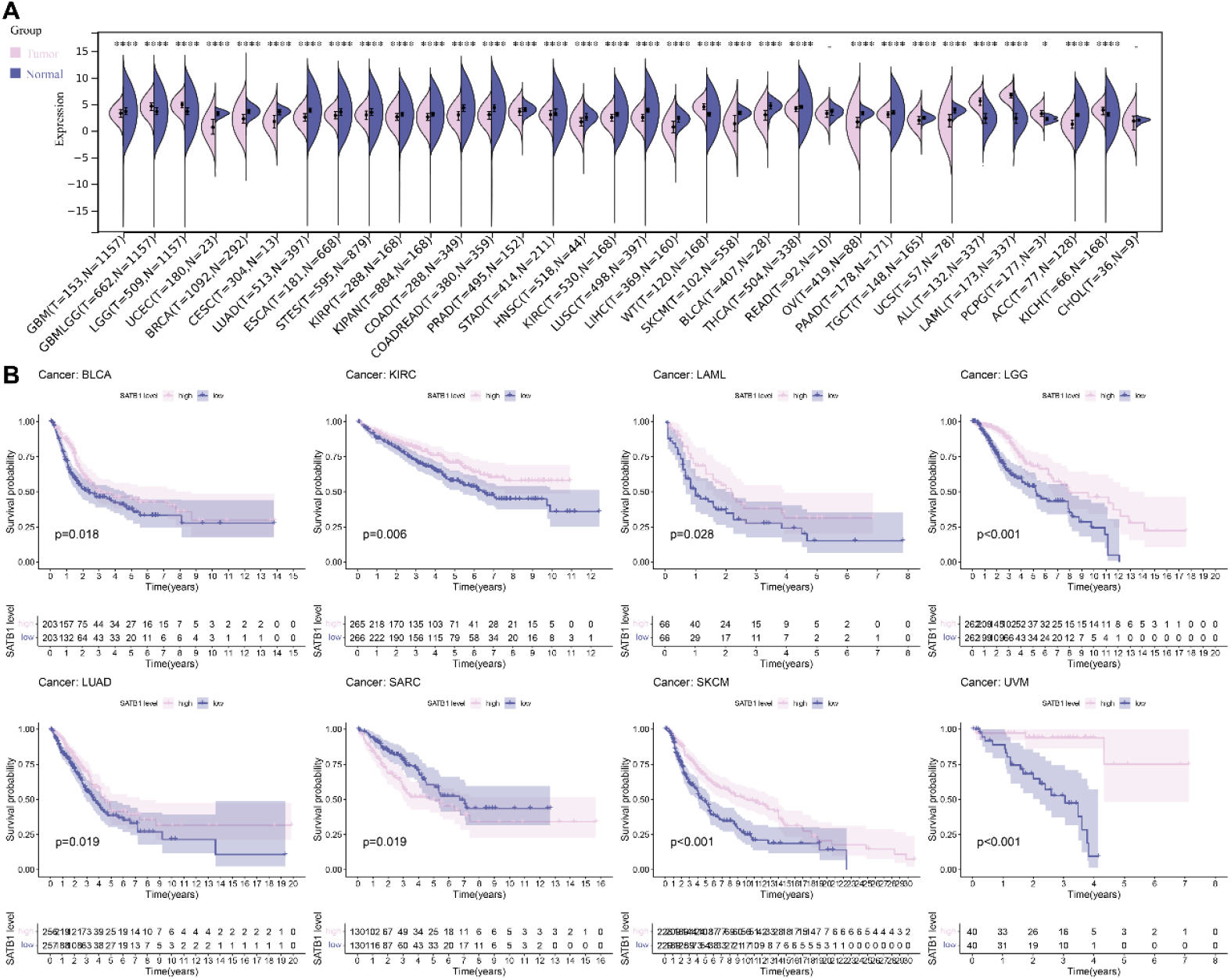
Pan-cancer analysis of SATB1-AS1. **A** Exploring the expression of SATB1-AS1 in pan-cancer. **B** The prognostic effect of SATB1-AS1 in other TCGA cancer types was investigated using UCSC Xena browser.

## 4 Discussion

Thymoma is a relatively inert tumor, but its pathogenesis is still unclear, and more features and targets are needed for early diagnosis, precise therapeutic targets, and prognosis prediction. The individualized treatment plan for THYM should be formulated by a multidisciplinary team of thoracic surgeons, medical oncologists, imaging physicians, pathologists, and radiotherapists on the basis of the patient’s TNM stage and Masaoka-Koga stage to provide optimal strategies for prolonging the patient’s long-term survival and reducing the side effects of treatment. The optimal strategy for prolonging long-term survival and minimizing the side effects of treatment should be developed by a multidisciplinary team of thoracic surgeons[3]. However, cancer treatment is limited by both intratumor and intertumor factors. However, cancer treatment is limited by intra- and intertumor heterogeneity, which requires the study of mechanisms that work across patient characteristics to develop innovative therapies.

In this study, we performed bioinformatics analysis in conjunction with data collected from publicly available databases with the aim of exploring eRNAs as prognostically relevant biomarkers in THYM. On the basis of the TCGA database, 134 eRNAs with prognostic value in THYM were identified. Among them, both SATB1-AS1 and the target gene SATB1 were strongly associated with patient prognosis. Low SATB1-AS1 and SATB1 expression was associated with poor clinical characteristics (stage staging). However, the role of SATB1-AS1 in prognosis was inconsistent across cancers. Low SATB1-

AS1 expression in BLCA, KIRC, LAML, LGG, LUAD, SARC, SKCM and UVM was associated with poor patient prognosis, suggesting that SATB1-AS1 may play a role as an oncogene in tumor progression. To investigate the role of SATB1-AS1 in the biological behavior of tumors in detail, GO enrichment analysis and KEGG pathway analysis of SATB1-AS1 were performed in this study. The results revealed that high expression of AP003469.2 was associated with the participation and regulation of leukocyte transendothelial migration, natural killer cell-mediated cytotoxicity, neutrophil extracellular trap formation, PD-L1 expression and the PD-1 checkpoint pathway in cancer, the T-cell receptor signaling pathway, Th1 and Th2 cell differentiation, Th17 cell differentiation and Th17 cell differentiation, and other molecules and pathways involved in cell biological behavior. It has been reported that tight regulation of the expression level of SATB1 isoforms and posttranslational modifications of the protein are essential for the regulatory role of SATB1 in T-cell development and that SATB1-deficient thymocytes are partially redirected to inappropriate T lineages and are unable to give rise to the NKT and Treg subpopulations[16, 17]. SATB1 is a key regulator of thymocyte development. Moreover, SATB1 is a genome organizer that is highly expressed in double-positive thymocytes, and SATB1 deletion leads to a variety of defects in T-cell development, including impaired positive and negative selection and impaired differentiation of thymic regulatory T cells (tTregs)[18, 19].

eRNAs act not only as tumor suppressors by regulating the expression of tumor suppressor genes but also as major regulators of oncogenes, exhibiting potential oncogenic functions[20]. The potential role of eRNAs in mediating cancer-related enhancer functions and gene transcription is also key to understanding how the rewiring of gene expression in cancer cells occurs[21]. eRNAs are also involved in immune checkpoint-associated pathways that regulate the immune response and influence tumor development[22]. In this study, we found that SATB1-AS1 was coexpressed with immune checkpoint-associated genes (e.g. CD-8, CCL5, and IFNG) in THYM, and in pancancer analyses, SATB1-AS1 was also coexpressed with immune checkpoint-associated genes enriched in pathways negatively regulating immune cell proliferation and activation in a variety of cancers. These analyses suggest that SATB1-AS1 may be a potential marker for the early diagnosis and prognosis of THYM and may be a promising immune-related therapeutic target for precision treatment of THYM patients. Although these studies provide new directions for exploring the mechanistic functions of eRNAs, many challenges remain: experiments are needed to further understand the functions and potential mechanisms of eRNAs in gene regulation and chromosomal interactions; the bioactivity of eRNAs and their correlation with diseases have not yet been fully elucidated; and, more importantly, the abundance of eRNAs in vivo is low and unstable, and more sensitive methods need to be developed to recognize eRNAs. More importantly, the abundance of eRNAs in vivo is low and unstable, and more sensitive methods need to be developed to recognize eRNAs.

A GWAS revealed that many disease-related genetic variants are enriched in enhancer elements, demonstrating the importance of eRNAs in the human genome from multiple perspectives[23]. The importance of eRNAs in the human genome has been demonstrated in many ways. Recent studies have identified wortmannin and valproic acid as potential anticancer drugs that can block eRNA activity, mainly by targeting major components of the extracellular matrix, including SPP1, COL1A1 and FN1[24]. Thus, eRNA blockade is a valuable area for further research in cancer therapy.

## 5 Conclusion

In this study, we explored the prognosis-related genes of THYM patients as well as the related target genes, discussing the possibility of eRNAs as potential biomarkers and disease therapeutic targets as well as some directions for future research. The results revealed the following: (1) SATB1-AS1 was identified for the first time as a highly positively correlated eRNA with its target gene SATB 1 among the prognosis-related eRNAs in THYM. (2) SATB1-AS1 may be an independent biomarker for the prognosis of THYM patients. (3) There was a significant correlation between SATB1-AS and immunomodulatory genes. (4) Differential expression of SATB1-AS1 across cancers and its impact on survival. Our findings may enhance the understanding of THYM tumorigenesis mechanisms, leading to better selection of therapeutic agents that individually target specific eRNAs and their cofactors.

## 6 Limitations

This study has several limitations. First, our study analyzed only the publicly available TCGA dataset and lacked a validation cohort to confirm the association between eRNA SATB1-AS1 and THYM prognosis. Second, although the relevance of the eRNA SATB1-AS1 having an immune checkpoint-associated gene was validated in pancancer, basic experiments are needed to confirm this relationship, which will be the focus of our follow-up study.

## Author Contributions

All authors contributed to the study conception and design. Material preparation, data collection and analysis were performed by Yichu Huang. The first draft of the manuscript was written by Hongpeng Wang and all authors commented on previous versions of the manuscript. All authors read and approved the final manuscript.

## Data availability

The datasets generated and/or analyzed in this study are available at The Cancer Genome Atlas (TCGA) database (https://portal.gdc.cancer.gov/).

